# The impact of Option B+ policy on engagement in HIV care and viral suppression among pregnant women living with HIV in South Africa

**DOI:** 10.1101/2024.11.08.24316965

**Authors:** Cornelius Nattey, Mhairi Maskew, Nelly Jinga, Amy Wise, Nicola van Dongen, Thalia Ferreira Brizido, Maanda Mudau, Karl-G Technau, Kate Clouse

**Author notes:** Equally contributing authors (Nattey and Maskew). **Corresponding author details:** Dr. Kate Clouse, Vanderbilt University School of Nursing. **Data availability statement:** Data may be obtained from a third party and are not publicly available. The maternal data underlying this article were provided with permission by the data gatekeeper for Rahima Moosa Mother and Child Hospital and the Empilweni Services and Research Unit. Cohort participants provided written consent for data to be used for research purposes, and requests for access can be directed to Empilweni Services and Research Unit, Johannesburg, South Africa. Laboratory data linked to the maternal cohort are owned by the National Health Laboratory Services and access is governed by policies and procedures in response to requests made directly to the NHLS Office of Academic Affairs and Research. As such, the corresponding author does not have authority to release the data to the public or other data-sharing repositories. However, these data can be requested by the public through standardized request forms, which are then considered in an internal review procedure.

## Abstract

**Background:** Early access to HIV care impacts maternal outcomes and the risk of vertical transmission of HIV Option B+, a policy that mandates offering all pregnant women living with HIV (PWLH) lifelong antiretroviral therapy (ART) irrespective of their CD4 count, has been adopted across sub-Saharan Africa, including South Africa since 2015. This study aimed to assess the impact of Option B+ on engagement in HIV care and viral suppression among pregnant women in South Africa.

**Methods:** This observational study used data from pregnant women living with HIV who delivered at Rahima Moosa Mother and Child Hospital in Johannesburg, South Africa from 2013-2017. Linkage to a national HIV laboratory cohort (the NHLS National HIV cohort) was used to ascertain engagement in HIV care prior to antenatal care (ANC) entry and viral load outcomes. Analyses were stratified by the pre-Option B+ (2013-2015) and Option B+ (2016-2017) eras. We compared engagement rates before and during the Option B+ era and assessed factors associated with HIV care engagement and viral suppression. Risk ratios were estimated using log-binomial regression. **Results:** Among 4,865 PWLH, 65% had evidence of prior engagement in HIV care. Prior engagement in care was higher during the Option B+ era (66%) compared to the pre-Option B+ era (55%) (p<0.001). Younger women (18-24 years) were less likely to engage in HIV care than those aged 25-34 years (aRR 0.8, 95% CI: 0.6-0.9). Women with CD4 counts <200 cells/mm³ were less likely to have been engaged in care prior to pregnancy compared to those with CD4 ≥500 (aRR 0.6, 95% CI: 0.6-0.7). Primigravid women had a 30% lower likelihood of earlier HIV care engagement compared to those with 2-3 pregnancies (aRR 0.7, 95% CI: 0.5-0.8). Overall viral suppression was higher in women reporting prior ART use compared to those with no prior HIV care (33% vs. 19%, p<0.001). During the four-year study period, the proportion of PWLH who had a viral load recorded but were not virally suppressed ranged from 22-36%.

**Conclusion:** The Option B+ policy led to increased engagement in HIV care prior to pregnancy. However, high prevalence of unsuppressed viral load during the Option B+ era highlights the need for continued monitoring and support to sustain the benefits of this policy. Pregnancy and antenatal care services remain an essential portal of entry to HIV care among PWLH in South Africa. Interventions to improve early ANC attendance and maternal engagement in HIV care prior to pregnancy are critical to eliminate vertical HIV transmission.

## INTRODUCTION

Option B+, a policy that mandates offering all pregnant women living with HIV (PWLH) initiate lifelong antiretroviral therapy (ART) irrespective of their CD4 count, has been adopted by most treatment programmes in sub-Saharan Africa [1], including South Africa in 2015 [2]. This policy aims to reduce the risk of vertical transmission of HIV through suppression of HIV viral load (VL) during pregnancy, while also potentially improving uptake and continuity of HIV care among women [3]. Option B+ adoption has dramatically increased the number of pregnant women initiating ART [4] and led to a substantial reduction in vertical transmission of HIV [5].

Though the implementation of Option B+ has also yielded high coverage of HIV testing at entry to antenatal care (ANC) [6], it is unclear whether it has affected the timing of engagement in HIV care in relation to ANC initiation. Engagement in HIV care prior to the start of ANC may reduce risk vertical transmission even further by ensuring complete ART coverage and sustained suppression of HIV viral load during pregnancy, delivery and the postpartum period [7–9]. Data from South Africa’s National Antenatal HIV Sentinel Survey indicate that, compared to women initiating ART before pregnancy, women who initiated ART during pregnancy achieved lower rates of viral load testing (73% vs. 83%) and viral suppression (57% vs. 76%) [10].

Despite the important potential benefits of early engagement in HIV care, many women still only learn their HIV status through routine antenatal testing and commencement of ART occurs late in pregnancy [11]. The impact of the implementation of Option B+ on engagement in HIV care and viral suppression at entry to antenatal care is unclear. Here, we estimate proportions of women with evidence of engagement in HIV care and viral suppression at entry to antenatal care. We also assess factors associated with engagement in HIV care prior to pregnancy. We then estimate the effect of prior engagement in HIV care on subsequent VL suppression during pregnancy and finally, stratify all analyses by vertical transmission policy era (prior to and during Option B+ era).

## METHODS

### Vertical transmission policies in South Africa

South Africa’s policy for providing antiretroviral therapy (ART) to pregnant women living with HIV (WLWH) has evolved over time, following World Health Organization (WHO) recommendations. In April 2008, a dual-therapy PMTCT programme was implemented through which women were offered Zidovudine starting at 28 weeks’ gestation or ART at a CD4 count of <200 cells/µL [12]. In 2010, Option A was introduced with Zidovudine starting at 14 weeks’ gestation for women with a CD4 count of >350 cells/µL or triple-drug ART for women with a CD4 count of ≤350 cells/µL [13]. The Option A regimen was replaced in 2013 by the WHO-recommended Option B approach, where women were offered triple-drug therapy throughout pregnancy, with postpartum withdrawal after cessation of breastfeeding for those ineligible for lifelong treatment [13]. The progression from Option B to Option B+ in 2015 was one of the key developments in the South African national HIV policy. Under Option B+, pregnant WLWH were offered lifetime ART regardless of CD4 count [2].

### Study population and data sources

This analysis uses two data sources. First, we used data from all pregnant women living with HIV who delivered at Rahima Moosa Mother and Child Hospital (RMMCH) in Johannesburg, South Africa, from 2013-2017. The RMMCH Cohort, supported by the Empilweni Services and Research Unit collects maternal and infant data including demographics, antenatal care, HIV treatment, delivery, early infant diagnosis of HIV, and maternal ART use [14].

Second, we used data from a national HIV cohort constructed using routine laboratory data from South Africa’s National Health Laboratory Service (NHLS), the sole provider of public sector laboratory services, serving 80-90% of the uninsured population. This cohort includes records of HIV-related tests used for treatment initiation and monitoring since 2004 [15]. Using an anonymized unique patient identifier previously developed and validated, individuals were followed longitudinally through their laboratory results as they progress through the HIV care and treatment cascade.

We used deterministic record linkage procedures using laboratory sample barcodes to link the RMMCH maternal cohort data to laboratory data from the NHLS National HIV Cohort. These alphanumeric barcodes are centrally allocated by NHLS and affixed to biological specimens by healthcare workers at the point of collection, usually clinics and hospitals. The barcode is the same across all the tests performed on the same person’s biological specimen and were captured in both NHLS and RMMCH databases. We manually validated a sample of linked data (1,200 records) comparing patient surname, first name and date of birth to ensure barcode linkages were correctly linking individuals across the two datasets. This process yielded exact matches for 87% of records and likely matches with minor typographical mismatches for a further 9% [16].

### Study variables

For this analysis, we defined prior engagement in HIV care at entry to ANC as either: 1) self-reported ART use prior to entry to antenatal care in maternal records; or 2) evidence of HIV-associated laboratory data (CD4 count, and viral load) observed in the NHLS national HIV cohort between three months prior to date of entry to antenatal care and 30 days after entry to antenatal care. The study period January 2013 to December 2015 was defined as being prior to Option B+ implementation, while January 2016 onwards was defined as the Option B+ era. Observed HIV viral load (VL) test results were classified into two groups: 1) VL suppressed was defined as an observed VL test result <400 copies/mL; 2) VL unsuppressed was defined as VL test results ≥400 copies/ml. We estimated VL outcomes at two time periods: 1) entry to antenatal care, which included any VL test within six months prior to date of entry to ANC and 2) the pregnancy period between date of entry to ANC and delivery date. We then considered if the implementation of the Option B+ policy modified the effect of prior engagement in care on VL outcomes by further stratifying these analyses by Option B+ era.

### Statistical analysis

We commenced the analysis by summarizing characteristics of mothers at entry to antenatal care using frequencies and simple proportions. Next, we summarised rates of engagement in HIV care and described characteristics of women with and without evidence of prior engagement in HIV care before the Option B+ era and during the Option B+ era. We then reported the frequency of VL testing at first antenatal care visit and during pregnancy. Among observed viral load tests, we summarise proportions with viral suppression by engagement in care prior to pregnancy or during pregnancy. In order to assess for effect measure modification of Option B+ era on effect of prior engagement in care on VL results at entry into ANC and during pregnancy, we stratified the proportion engaged in care prior to pregnancy achieving viral load suppression by Option B+ policy era. Lastly, we used log-binomial regression to report crude and adjusted relative risks (aRR) and 95% confidence intervals (95%CI) of factors associated with engagement in HIV care prior to pregnancy and viral load suppression after adjusting for potential confounders.

### Ethical considerations

This study was approved by the Human Research Ethics Committee (HREC) of the University of Witwatersrand. This is a secondary analysis of de-identified data collected as part of routine care and no direct participant interaction occurred.

## RESULTS

### Characteristics of study participants at entry to antenatal care

A total of 4,865 pregnant women living with HIV delivered infants at RMMCH during the study period 2013-2017 and were included in analyses (**Figure 1)**.

**Figure 1:**
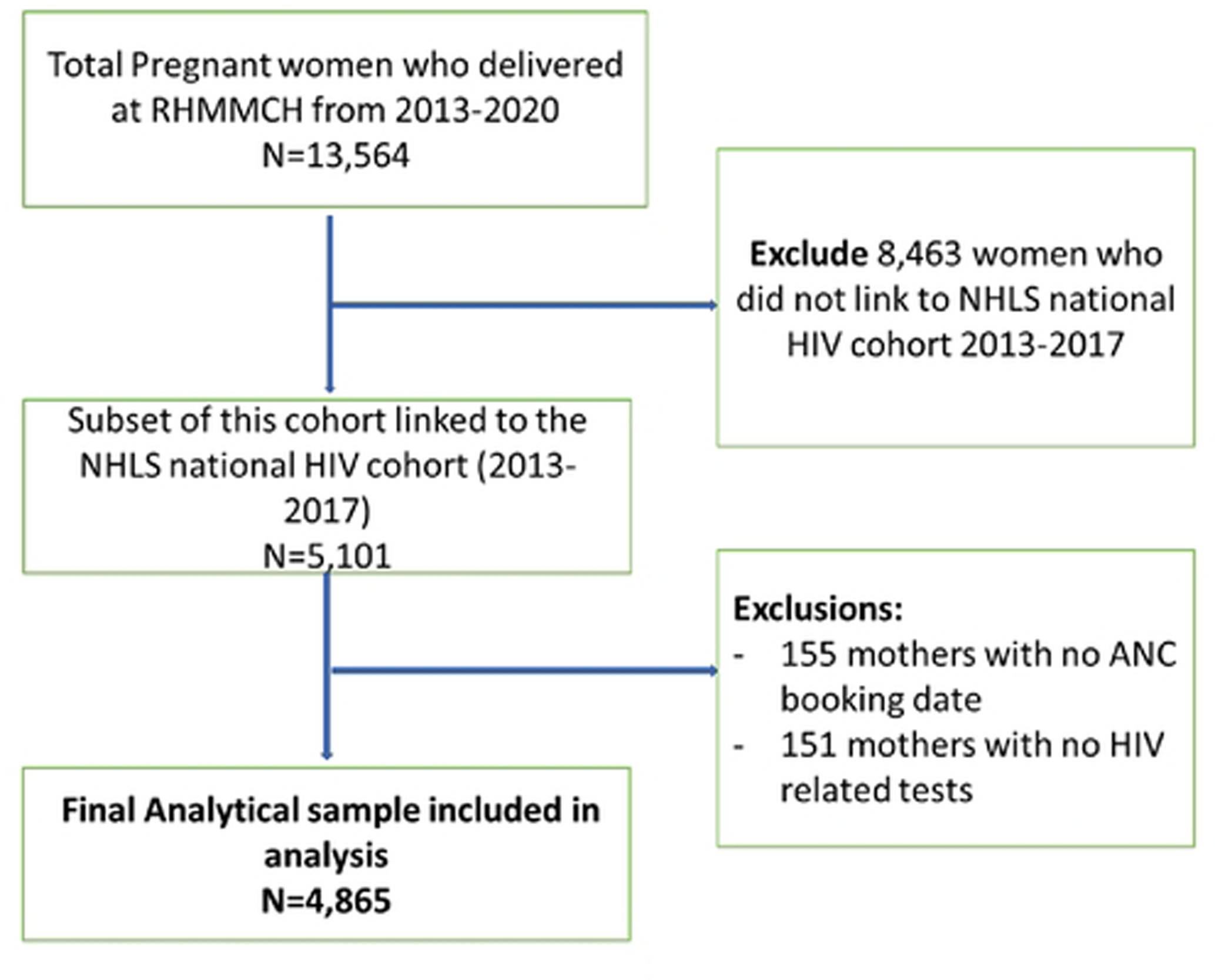
Study eligibility flow chart of cohort of pregnant women living with HIV delivered infants at RMMCH during the study period 2013-2017

Participant characteristics are shown in **Table 1**. The median maternal age at entry to antenatal care was 31 (IQR:26-35) years and median gestational age at entry to antenatal care was 21 (IQR:15-26) weeks. Overall, the median CD4 count cell count was 393 cells/mm^3^ (IQR:252-554). A total of 17.0% of women had CD4 counts <200 cells/mm³, signifying advanced stages of HIV infection and considerable immune system compromise. Stratification by gestational age at entry to ANC revealed that 11% of women booking in the first trimester had CD4 counts <200 cells/mm³, increasing to 18% in the second trimester and 19% in the third trimester. At first ANC visit, approximately half (51%) of women were in their second trimester while >80% had at least one other child. The majority of mothers were initiated onto standard first line as efavirenz based ART regimens with tenofovir and either lamivudine or emtricitabine. ART regimens with efavirenz and either lamivudine or emtricitabine (88%).

**Table 1:**
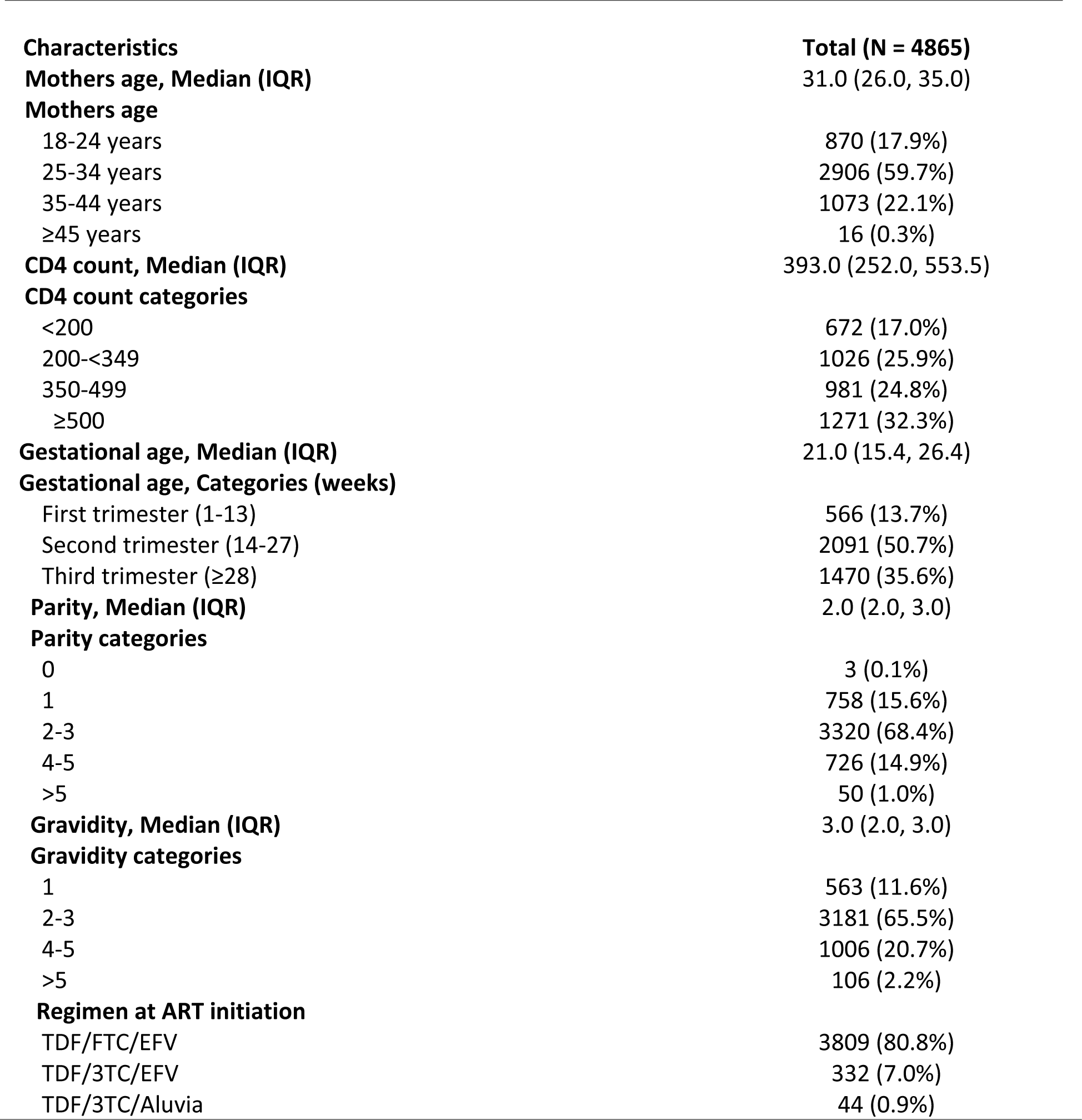
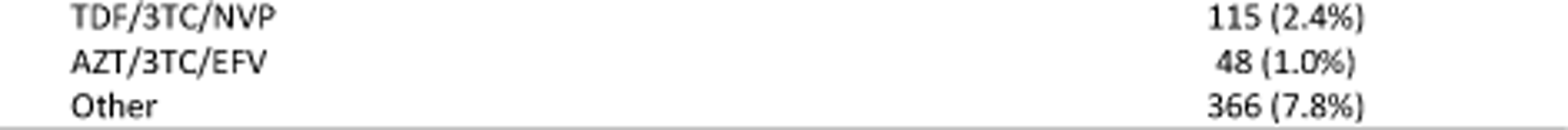
Characteristics of study participants at entry to antenatal care (n=4,865).

| **Characteristics** | **Total (N = 4865)** |
| --- | --- |
| **Mothers age, Median (IQR)** | 31.0 (26.0, 35.0) |
| **Mothers age** |  |
| 18-24 years | 870 (17.9%) |
| 25-34 years | 2906 (59.7%) |
| 35-44 years | 1073 (22.1%) |
| ≥45 years | 16 (0.3%) |
| **CD4 count, Median (IQR)** | 393.0 (252.0, 553.5) |
| **CD4 count categories** |  |
| <200 | 672 (17.0%) |
| 200-<349 | 1026 (25.9%) |
| 350-499 | 981 (24.8%) |
| ≥500 | 1271 (32.3%) |
| **Gestational age, Median (IQR)** | 21.0 (15.4, 26.4) |
| **Gestational age, Categories (weeks)** |  |
| First trimester (1-13) | 566 (13.7%) |
| Second trimester (14-27) | 2091 (50.7%) |
| Third trimester (≥28) | 1470 (35.6%) |
| **Parity, Median (IQR)** | 2.0 (2.0, 3.0) |
| **Parity categories** |  |
| 0 | 3 (0.1%) |
| 1 | 758 (15.6%) |
| 2-3 | 3320 (68.4%) |
| 4-5 | 726 (14.9%) |
| >5 | 50 (1.0%) |
| **Gravidity, Median (IQR)** | 3.0 (2.0, 3.0) |
| **Gravidity categories** |  |
| 1 | 563 (11.6%) |
| 2-3 | 3181 (65.5%) |
| 4-5 | 1006 (20.7%) |
| >5 | 106 (2.2%) |
| **Regimen at ART initiation** |  |
| TDF/FTC/EFV | 3809 (80.8%) |
| TDF/3TC/EFV | 332 (7.0%) |
| TDF/3TC/Aluvia | 44 (0.9%) |
| TDF/3TC/NVP | 115 (2.4%) |
| AZT/3TC/EFV | 48 (1.0%) |
| Other | 366 (7.8%) |

### Engagement in HIV care prior to entry to antenatal care

Overall, we found evidence of prior engagement in HIV care before entry to ANC for 65% of women (n=3148). Of these, 10% (315/3148) self-reported prior ART use, 39% (1223/3148) had laboratory evidence of prior ART and 51% (1610/3148) had both. **Figure 2** depicts trends in engagement in HIV care from 2013-2017. Overall, we found increasing evidence of engagement in HIV care prior to pregnancy by year through the study period, particularly among multigravida women. Proportions with prior engagement in HIV care increased from 17% in 2013 to 53% in 2017 among primigravid women and from 49% in 2013 to 73% in 2017 among multigravida women. More women engaged in HIV care before the first ANC visit during Option B+ era, compared to the period prior to Option B+ implementation (66% (2803/4,241) versus 55% (345/624), respectively).

**Figure 2:**
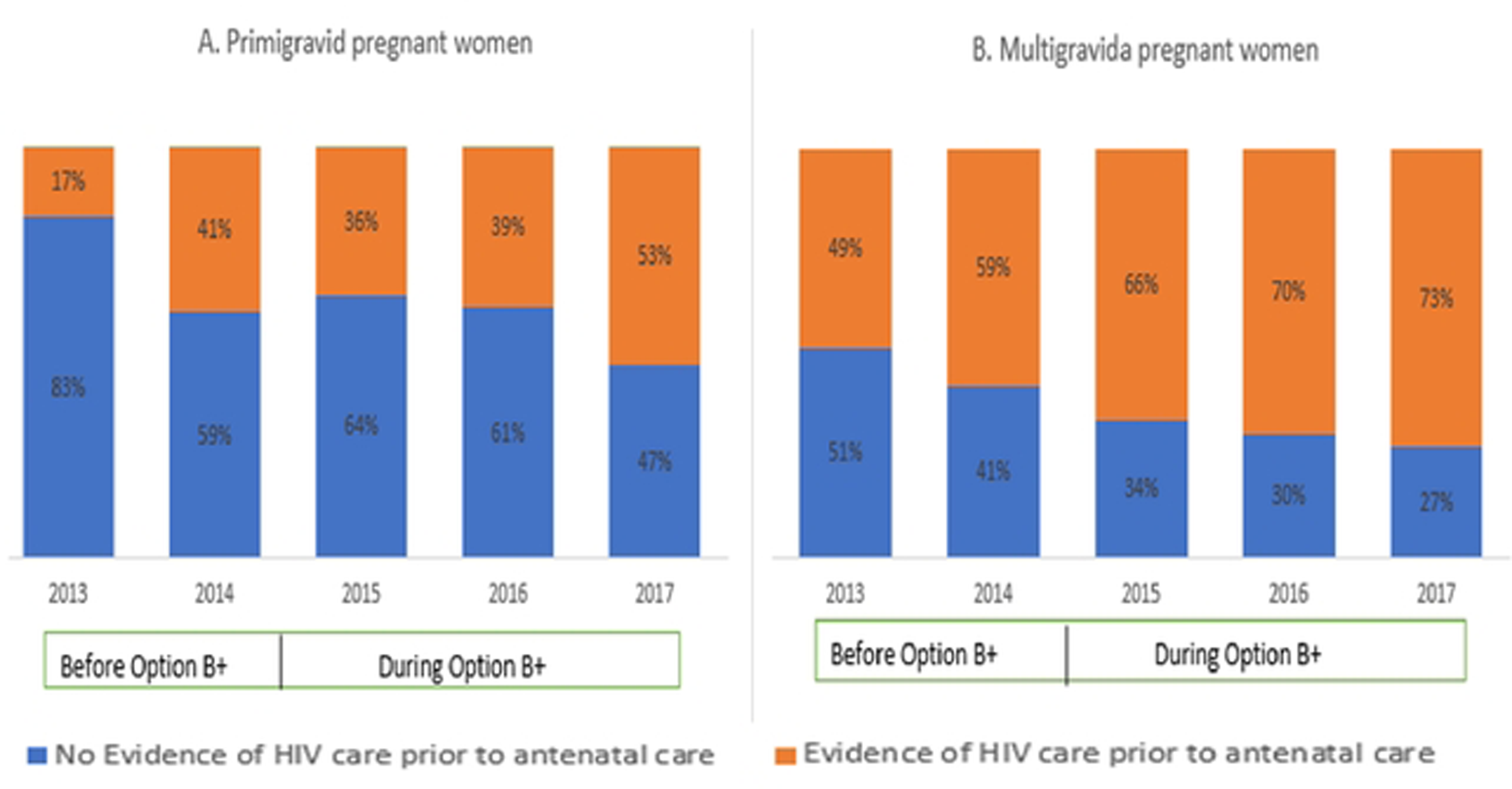
Engagement in HIV care before and during Option B+ era among (A) primigravid pregnant women and (B) multigravida pregnant women

Factors associated with engagement in HIV care among pregnant women accessing ANC services are summarized in **Table 2**. Younger women (age 18-24) were 20% less likely to have evidence of HIV care engagement prior to ANC compared to those 25-34 years (aRR 0.8; 95% CI:0.5-0.9).

**Table 2:**
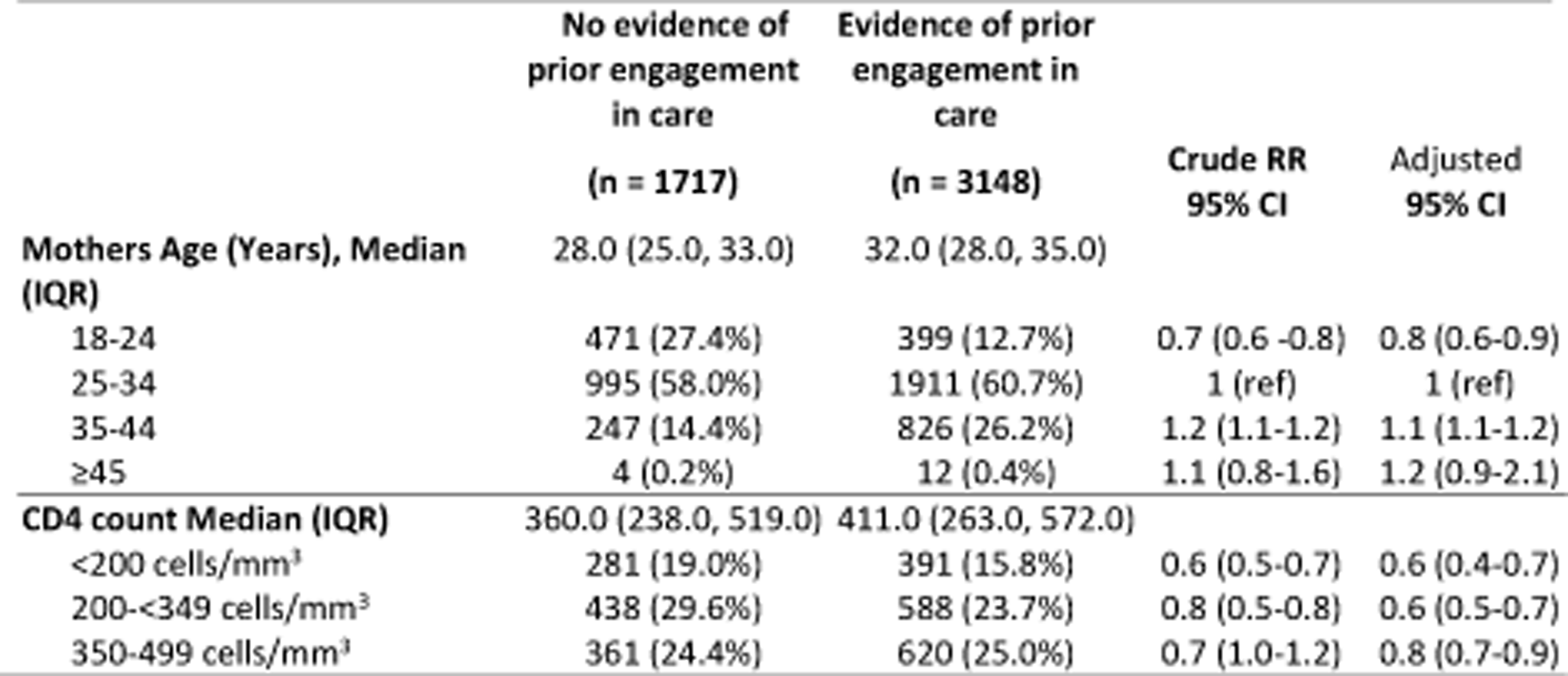
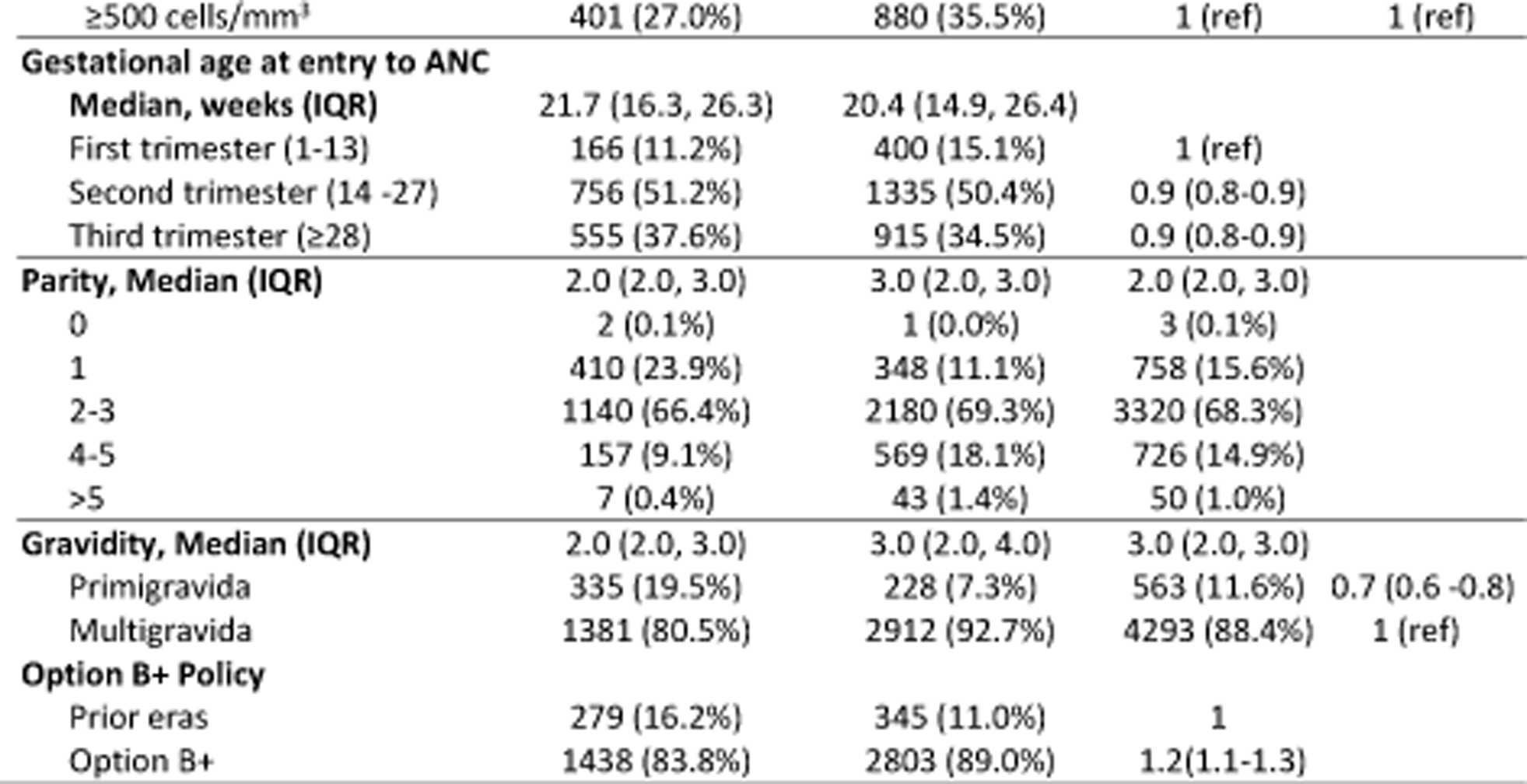
Engagement in HIV care prior to antenatal care and predictors of prior engagement in HIV care.

Those with very low CD4 (<200 cells/mm^3^) were 40% less likely to have evidence of earlier HIV care, compared to those with a CD4 of ≥500 cells/mm^3^ (aRR 0.6; 95% CI: 0.6-0.9). Women who were primigravid were also less likely to engage in HIV care early, compared to those with 2-3 pregnancies, (aRR: 0.7; 95%CI: 0.4-0.6).

### HIV Viral load monitoring and viral suppression

A total of 4,910 viral loads were observed between entry to ANC and delivery among the 4,865 study participants. Of these, 41% (2002/4865) women had a VL test at entry to ANC, and 60% (2908/4865) had at least one VL observed between ANC booking date and delivery date.

Overall, 48% (n=1520) of the 3148 women with evidence of engagement in prior HIV care had an observed VL at entry to ANC (**Figure 3A**). Of these, 61% (922/1520) were virally suppressed. In contrast, of the 1717 women with no evidence of prior HIV care at ANC, 482 (28%) had a VL test at entry to ANC and 32% (153/482) of these were virally suppressed. Overall, viral suppression at ANC was higher among pregnant women reporting prior ART use compared to women reporting no HIV care prior to entry into ANC 29% (922/3148) versus 9% (153/1717) suppressed. Overall population level VL suppression at ANC was 22% (1075/4865).

**Figures 3A and 3B:**
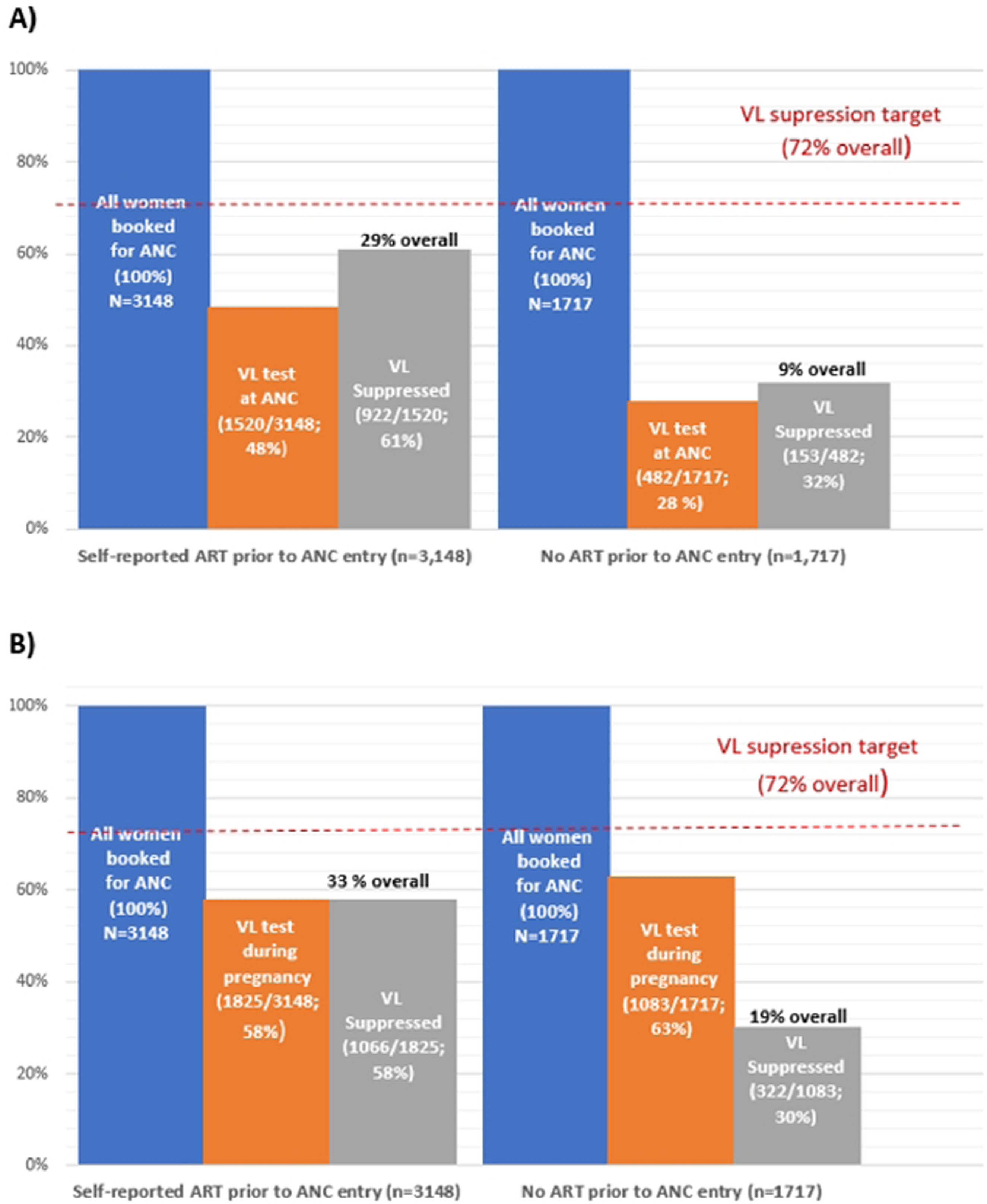
HIV viral suppression at A) entry to ANC and B) during pregnancy stratified by use of ART prior to ANC entry

During pregnancy, VL testing was observed for 58% (n=1825) of the 3148 women with evidence of engagement in prior HIV care (**Figure 3B**). Of these, 58% (1066/1825) were virally suppressed. Among the 1717 women with no evidence of prior HIV care at ANC, 1083 (63%) had a VL test at entry to ANC and 30% (322/1083) of these were virally suppressed. Overall, rates of viral suppression during pregnancy were higher among pregnant women reporting prior ART use compared to suppression rates observed among women reporting no HIV care prior to entry into ANC (33% (1066/3148) vs 19% (322/1717) suppressed). Overall population level VL suppression during pregnancy was 29% (1388/4865).

**Figure 4** represents trends in viral suppression among women from 2014 to 2017 during pregnancy. In 2014 and 2015, a substantial proportion had suppressed VL (76% and 78%, respectively). However, in 2016, there was a notable increase in unsuppressed VL, rising to 36%; there was a slight recovery in 2017, with 67% suppressed, though was still higher than 2014 and 2015.

**Figure 4:**
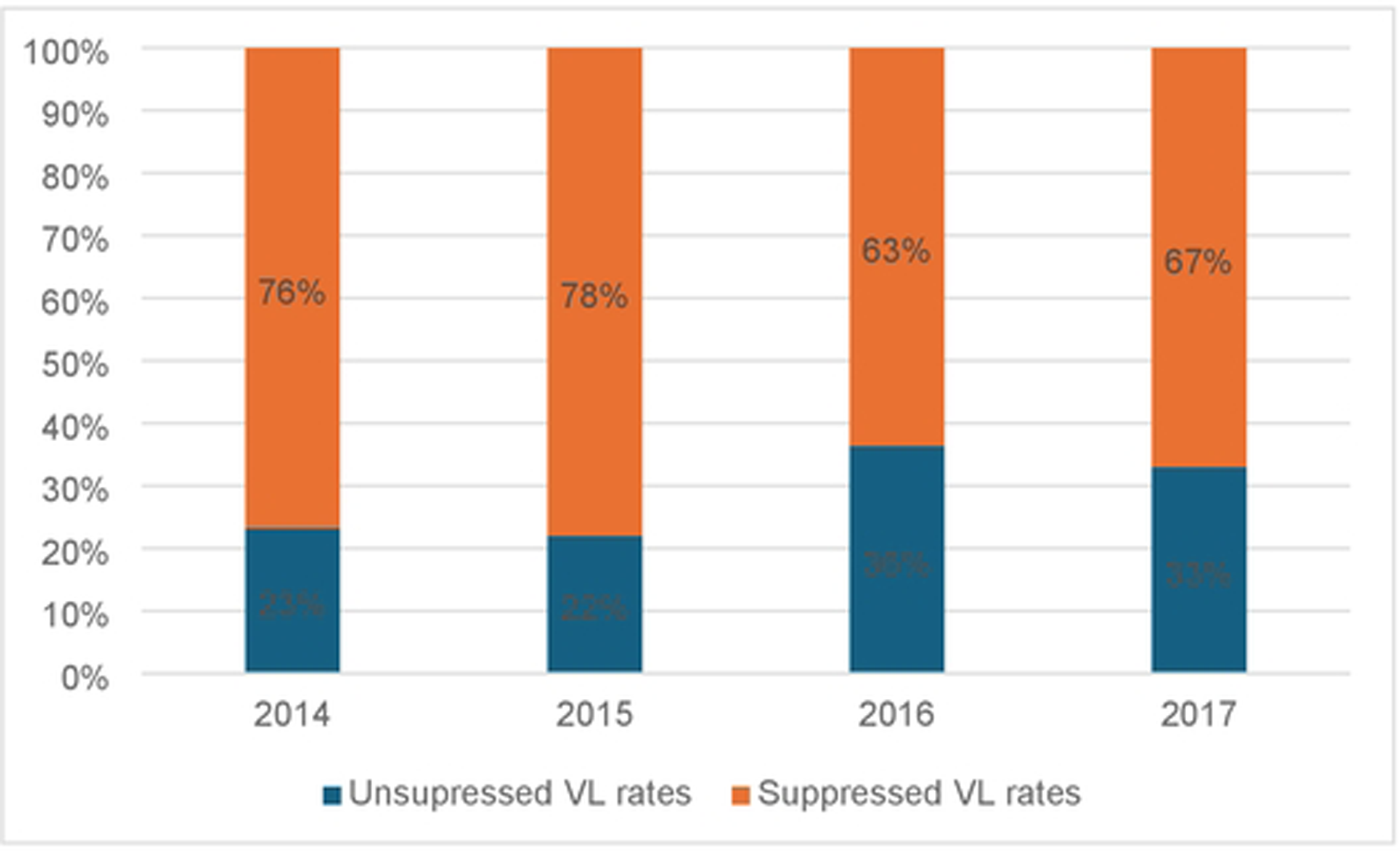
Trends in viral load suppression rates during pregnancy, 2014-2017

Factors associated with viral load suppression between ANC until delivery from multivariate log-binomial regression models are presented in **Table 3**. Women who started ART during ANC were less likely to achieve viral suppression than those who reported starting ART prior to pregnancy (aRR 0.9; 95% CI:0.8-0.9). Women initiating ART with higher CD4 count (≥500 vs <200) were more likely to achieve viral load suppression (aRR 1.6; 95% CI:1.4-1.7).

**Table 3:**
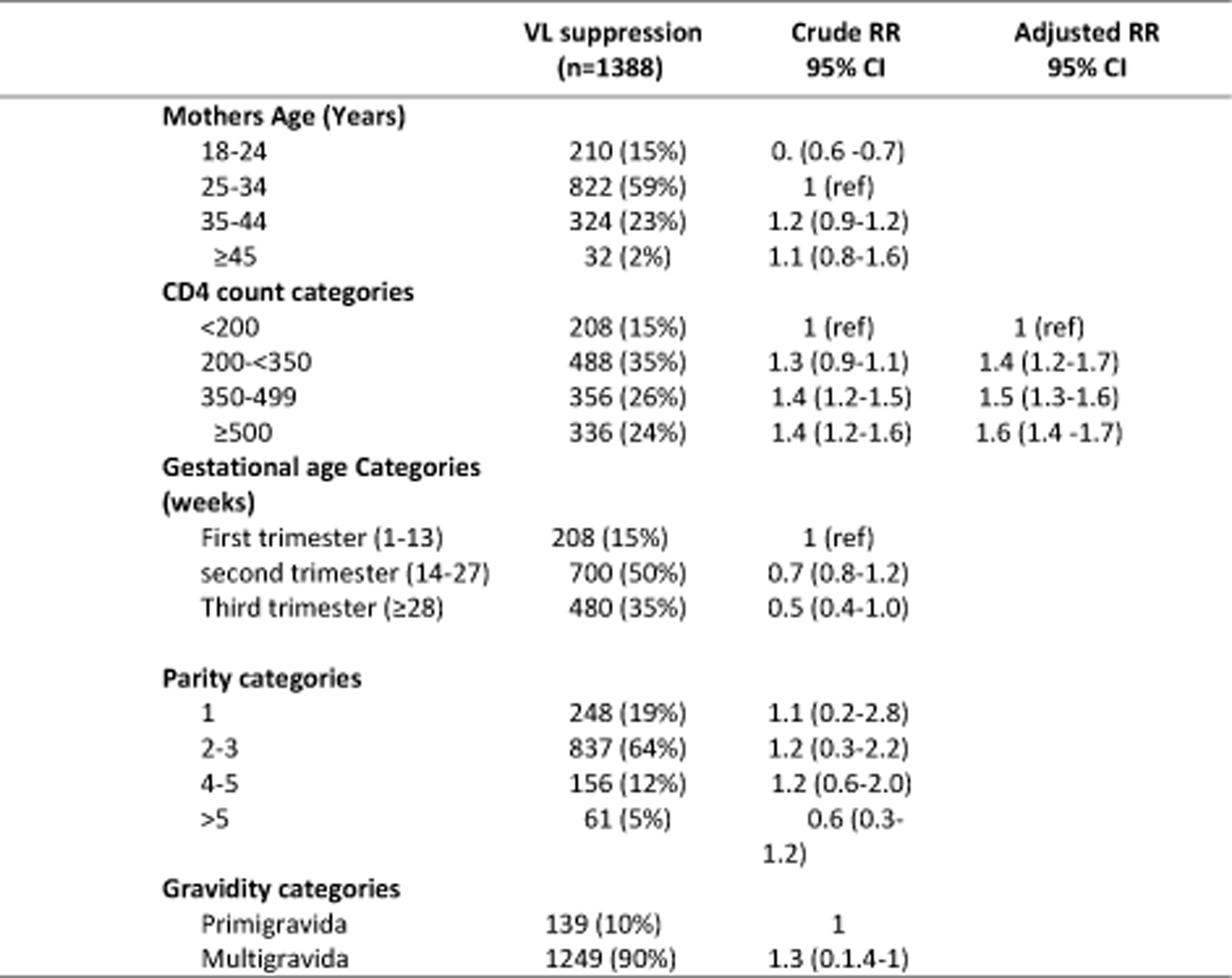
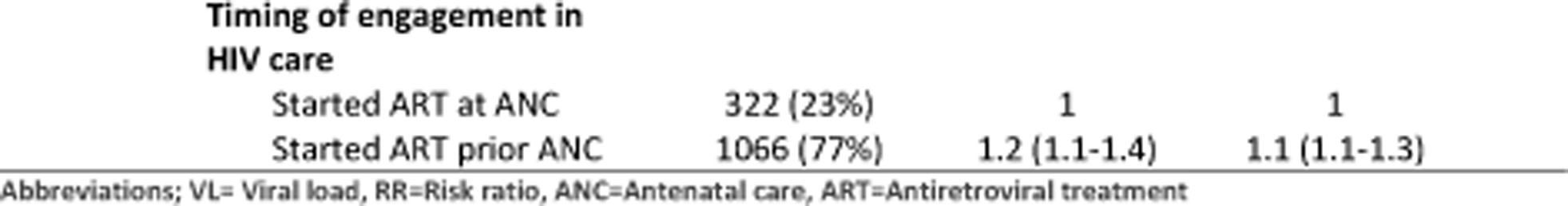
Multivariate log-binomial regression model of factors associated with viral suppression among pregnant women during pregnancy.

Engagement in HIV care prior to entry to ANC was associated with a higher probability of viral suppression both before and after the implementation of Option B+, with the effect being statistically significant except for those ANC initiation before Option B+, as shown in **Table 4**. The introduction of Option B+ seems to have improved the effectiveness of engagement in prior care on VL suppression outcomes, especially noticeable with a narrow confidence interval post-intervention, indicating more precise estimates of the effect.

**Table 4:** Effect measure modification analysis of the impact of Option B+ era on effect of prior engagement in care on VL results at entry to ANC and during pregnancy.

| At ANC initiation |  |  | During Pregnancy |  |  |
| --- | --- | --- | --- | --- | --- |
| Engagement in Prior Care | VL suppression | VL Risk Ratio | Engagement in Prior Care | VL suppression | Risk Ratio |
| **Before Option B+** |  |  |  |  |  |
| **N=79** | **N=51** |  | **N=184** | **N=142** |  |
| No (n=19) | 9 (17%) | 1 | No (n=56) | 39 (27%) | 1 |
| Yes (n=60) | 42 (83%) | 1.41 (0.79-2.51) | Yes (n=128) | 103 (73%) | 1.31 (1.02-1.69) |
| **After initiation of Option B+** |  |  |  |  |  |
| **N=1,923** | **N=1,312** |  | **N=2,724** | **N=1,876** |  |
| No (n=463) | 244 (19%) | 1 | No (n=752) | 488 (26%) | 1 |
| Yes (n=1460) | 1,068 (81%) | 1.39 (1.20-1.50) | Yes (n=1,972) | 1,388 (74%) | 1.09 (1.01-1.18) |

## Discussion

Universal access to HIV care and treatment has been a reality for pregnant women in South Africa for over two decades [17]. Despite this, engagement in HIV care remains below treatment targets [18]. This observational study describes the engagement of pregnant women living with HIV in care prior to pregnancy and its impact on viral load monitoring and suppression, particularly in the context of changing HIV care policies. We report several key findings concerning engagement in HIV care prior to pregnancy. First, despite expanded access to HIV care, antenatal services continue to be a critical entry point for HIV care. In 2017, nearly half of primigravida women had no prior exposure to HIV care at their ANC booking, posing significant risks to their health and increasing the likelihood of vertical transmission to their infants. However, the expansion of access to ART as implemented under the Option B+ policy appears to have had a positive impact on engagement in care. More women (66%) engaged in HIV care before the first ANC visit during the Option B+ era, compared to 55% before Option B+. Similar improvements in engagement in care have been observed in the region including a Malawian study that reported more women living with HIV entering ANC already on ART after introduction of Option B+ (18.7% pre-versus 30.2% post-Option B+) compared to the era prior to Option B+ implementation [19]. Data from Zambia also showed an increase in those on ART at first ANC visit from 9% in 2011, before Option B+, to 74% in 2015 during Option B+ [20]. Timely engagement in ANC may influenced not only by structural barriers in terms of access to care but also dissemination of information about timely HIV care [21].

Secondly, early access to ART remains a critical step towards eliminating vertical transmission of HIV [18], as sustained viral suppression through continuous ART use is essential for this goal. Our findings highlight that increased maternal engagement in HIV care prior to pregnancy, particularly through early ART initiation, is crucial. We found that pregnant women who reported prior ART use had increased uptake of viral load testing and a higher proportion of viral suppression during pregnancy compared to those who initiated ART during pregnancy. Hence, interventions are needed to improve maternal engagement in HIV care before pregnancy to ensure sustained viral suppression and reduce the risk of vertical transmission. This findings are consistent with a previous study among participants initiating ART prior to pregnancy having had higher viral suppression [22].

Thirdly, we found that maternal age is an important factor in patterns of engagement in HIV and antenatal care. Younger women in our study were less likely (20%) to engage in HIV care before ANC compared to those aged 26-34 and older, consistent with other work indicating younger maternal age is a risk factor for poor HIV care engagement [10]. This pattern is closely linked with parity, as younger women are more likely to be experiencing their first pregnancy and thus new to both HIV care and ANC services. By the second pregnancy, rates of engagement in HIV care prior to ANC improve, as maternal age increases and women may have already been exposed to HIV care during a prior pregnancy [23]. Therefore, age on its own is not the sole driver of accessing care, but is influenced by prior exposure to care through earlier pregnancies.

Barriers to maternal HIV care engagement identified among younger mothers in other studies include instability in romantic relationships [24,25], difficulty accepting one’s HIV status [26], and fear of beginning lifelong treatment [27]. Efforts to support younger women in maternal HIV care should focus on strengthening social support networks, either by engaging existing supporters or introducing external peer supporters, such as other young women living healthy lives with HIV. Similarly, primigravid women were less likely to engage in prior HIV care compared to multigravida women. A recent study among women with an established HIV diagnosis at ANC found that those pregnant for the first time had an increased risk of poor care engagement, consistent with our findings [10]. This may be attributed to a lack of knowledge or experience with HIV testing and care services, fear of stigma or discrimination, and limited access to services [28,29]. Additionally, pregnancy and the postpartum period are high-risk times for HIV acquisition, further complicating engagement in care.

Despite continually expanding access to ART for pregnant women, we noted persistence of advanced HIV disease at presentation. Overall 17% of women entering antenatal care did so with a CD4 count <200 cells/mm^3^. Unsurprisingly, those with low CD4 at entry to care were less likely to have engaged in HIV care prior to the current pregnancy. A previous study of a retrospective cohort of adult women also found that those with pre-pregnancy ART were more likely to start ANC with CD4 count ≥500 [24]. Pre-pregnancy ART improves immunologic and virologic control during pregnancy and there is therefore a need for renewed efforts in HIV testing, linkage to ART and viral monitoring. Additionally, stratification by gestational age at entry to ANC revealed concerning trends: 11% of women booking in the first trimester had a CD4 count <200 cells/mm^3^, while this proportion increased to 18% in the second trimester and 19% in the third trimester. These findings suggest that delayed ANC booking is associated with a higher likelihood of presenting with advanced immunosuppression. Thus, there is a pressing need to promote early ANC attendance, especially for women not previously engaged in HIV care, to reduce the burden of advanced HIV disease.

Finally, we noted with concern, low rates of VL testing during pregnancy (58%). Though this estimate is higher than previous estimates reported 20% in Gauteng Province, South Africa [25], 30% in Mozambique [30], and 40% from three districts of Kwazulu-Natal Province, South Africa [26], low rates VL monitoring during pregnancy calls for renewed attention to VL testing efforts during ANC. Our overall population level VL suppression during pregnancy of 29% is substantially lower than the UNAIDS target of 73%. While increased monitoring is important, improving VL suppression rates requires enhanced adherence support, timely ART initiation before pregnancy, and targeted interventions to address barriers to sustained ART use among pregnant women [27]. The finding that women with higher CD4 counts at the start of ART had a 1.5 times higher likelihood of achieving viral load suppression is consistent with the well-established benefits of initiating ART early in the course of HIV infection [31]. This result underscores the importance of early diagnosis and linkage to care, which allows for timely initiation of ART. By starting treatment at a higher CD4 count, the immune system is better preserved, and HIV can be suppressed more effectively. This not only improves health outcomes but also reduces the risk of transmission to others.

Our results should be interpreted in light of some limitations. Firstly, linking of maternal records to the National HIV cohort restricted our analysis to 2013-2017 since the NHLS cohort data are available until March 2018 but maternal records from RMMCH are through 2021. Secondly, while linking maternal and laboratory test datasets offers the opportunity to analyse laboratory results for ART clients beyond their originating facility, increasing the robustness of our estimates for VL testing, there are still challenges related to the generalizability of our findings. The cohort primarily reflects the population and practices of a single facility, which may not be representative of all ART clients in different regions or settings, especially outside urban areas. Moreover, missing data, particularly in the form of incomplete records or discrepancies between datasets, may introduce bias. This could potentially affect the accuracy of our estimates and should be considered when interpreting the findings.

Despite these limitations, our study leverages linking of maternal and laboratory test datasets which offers the opportunity to analyse laboratory results for ART clients beyond their originating facility, increasing the robustness of estimates for VL testing.

## Conclusion

Expanded access to HIV care and treatment for pregnant women living with HIV under the Option B+ policy appears to have increased rates of engagement in HIV care prior to entry to antenatal care. Despite this, pregnancy and antenatal care services remain an essential portal of entry to HIV care among women living with HIV in South Africa, and a large proportion of pregnant women living with HIV still present for HIV care with advanced HIV disease. Prior use of ART at entry to antenatal care is also associated with other positive treatment outcomes including timely monitoring of HIV viral load and also likelihood of viral suppression during pregnancy.

## Data Availability

Data may be obtained from a third party and are not publicly available. The maternal data underlying this article were provided with permission by the data gatekeeper for Rahima Moosa Mother and Child Hospital and the Empilweni Services and Research Unit. Cohort participants provided written consent for data to be used for research purposes, and requests for access can be directed to Empilweni Services and Research Unit, Johannesburg, South Africa. Laboratory data linked to the maternal cohort are owned by the National Health Laboratory Services and access is governed by policies and procedures in response to requests made directly to the NHLS Office of Academic Affairs and Research. As such, the corresponding author does not have authority to release the data to the public or other data-sharing repositories. However, these data can be requested by the public through standardized request forms, which are then considered in an internal review procedure.

## Acknowledgements

We wish to acknowledge the research study teams at Rahima Moosa Mother and Child Hospital and the Empilweni Services and Research Unit who collected data on all cohort participants and provided on-going data quality support and access. We also wish to acknowledge the South African National Health Laboratory Services for access to laboratory data.

## Notes

**Funding:** This study was funded by the US National Institutes of Health (NIH) Eunice Kennedy Shriver National Institute of Child Health & Human Development and the National Institute for Allergy and Infectious Diseases under grant R01HD103466 and R01HD103466-04S1. The RMMCH cohort was also supported by the Eunice Kennedy Shriver National Institute of Child Health and Human Development and the National Institute of Allergy and Infectious Disease under grant U01HD080441 and U01AI069924. The views expressed are solely those of the authors and do not necessarily represent the views of the NIH. The funding source had no role in the design of this study and did not have any role during its execution, analyses, interpretation of the data or decision to submit the results.

### Competing Interest Statement

The authors have declared no competing interest.

### Funding Statement

Yes

### Author Declarations

Analyses of data from the Rahima Moosa Mother and Child Hospital Maternal HIV Cohort and linkage of this cohort to laboratory data from the National Health Laboratory Services approved under protocol M200237 of the Human Research Ethics Committee (Medical) of the University of the Witwatersrand and the IRB of Boston University.

